# Establishing Thrombolysis in Myocardial Infarction Frame Count as a Clinical Measure of Coronary Microvascular Disease in Women

**DOI:** 10.1101/2022.09.24.22280321

**Authors:** Nicole Wayne, Qufei Wu, Stephen C. Moore, Victor A. Ferrari, Scott D. Metzler, Marie A. Guerraty

## Abstract

**Background:** Despite growing awareness of the crucial role of the coronary vasculature in cardiovascular health and disease, diagnosing coronary microvascular disease (CMVD) remains challenging because it often requires advanced cardiac imaging that are available at only a few centers. For example, perfusion positron emission tomography (PET) allows for the quantification of the myocardial blood flow (MBF) and, thus, the calculation of the MBF reserve (MBFR), which is the ratio of MBF under rest conditions and MBF under hyperemic or stress conditions. In the absence of obstructive coronary artery disease, MBFR is a measure of CMVD. However, the availability of perfusion PET is limited. Thrombolysis in Myocardial Infarction (TIMI) frame count (TFC) is an angiography-based measure of coronary flow that has been proposed, but not well established, as a measure of CMVD. We aim to demonstrate a relationship between TFC and MBFR and to establish cut-off measures from clinical coronary angiograms for the diagnosis of CMVD.

**Methods:** We identified a cohort of 123 patients (age 58 +/− 12.1, 63% female, 41% Caucasian) who had no obstructive coronary artery disease and had undergone perfusion PET stress testing and clinical coronary angiography for clinical indications. We compared TFC for each coronary territory with regional perfusion PET parameters using linear regression modeling. We then used two mathematical models of the coronary circulation to understand the relationship between these parameters. We performed ROC analysis to determine the ability of TFC to diagnose CMVD, defined as global MBFR < 2.

**Results:** There is a very tight sex-dependent correlation between TFC and MBFR, but no association between resting coronary flow and TFC and only a weak association between stress coronary flow and TFC. Mathematical modeling of the coronary circulation highlights an uncoupling between TFC and flow in larger vessels where TFC is measured, providing a likely explanation for the surprising empiric result. ROC analysis shows TFC as an excellent measure of CMVD in women (AUC 0.84-0.89) and establishes TFC cutoffs.

**Conclusions:** TFC from clinical coronary angiograms reflects coronary microvascular function in a sex-dependent manner and performs well in identifying women with CMVD.

**Clinical Perspective:** *What is new?:* Thrombolysis in Myocardial Infarction (TIMI) frame count (TFC) is highly correlated with myocardial blood flow reserve (MBFR), which reflects coronary vasodilatory potential, in patients without obstructive coronary artery disease. Computational modeling establishes how the relationship between TFC and MBFR can exist. TFC performs much better in women than in men for the diagnosis of coronary microvascular disease (CMVD) and thresholds for the diagnosis of CMVD with TFC are identified.

*What are the clinical implications?:* TFC from clinical coronary angiograms can be used to assess CMVD in women.

## Introduction

Coronary microvascular disease (CMVD) leads to angina, myocardial infarction, and heart failure ^1, 2^ and accounts for 30-50% of the burden of ischemic heart disease (IHD). CMVD also worsens the prognosis in patients with coronary artery disease (CAD) and is an independent risk factor for major adverse cardiovascular events^3, 4^. However, challenges in diagnosing CMVD have limited both CMVD research and clinical care of individuals with suspected or confirmed CMVD. Furthermore, although the prevalence of CMVD is similar in men and in women^5^, the underdiagnosis of CMVD in women contributes to the discrepant burdens of untreated heart disease between women and men^1, 2, 6, 7^.

Recent advances in imaging technology have allowed researchers to study and diagnose IHD due to CMVD. Positron emission tomography (PET) perfusion imaging allows for the quantification of myocardial blood flow (MBF) and MBF reserve (MBFR). MBFR is a unitless ratio between MBF during hyperemic or stress conditions and rest MBF that reflects coronary microvascular function, or the ability of the coronary microvasculature to vasodilate appropriately. In the absence of epicardial coronary artery disease, these measures directly reflect coronary microvascular health. PET is the current gold standard for non-invasive diagnosis of CMVD, but its availability is limited to selected tertiary care centers^8^. Other imaging modalities to diagnose CMVD are actively being studied and developed, including magnetic resonance imaging and computed tomography (CT)-based techniques^8, 9^.

In addition to advanced cardiac imaging, a method based on coronary flow during angiography – termed Thrombolysis in Myocardial Infarction (TIMI) frame count (TFC) – has been described^10^. TFC estimates the amount of time that contrast material takes to fill the epicardial vessels during a coronary angiogram. Depending on the dose and type of sedation used during the coronary angiogram, TFC may measure a rest state or a hyperemic state. High TFC is associated with worse prognosis in the context of myocardial infarction^10, 11^ and has also been shown to be an independent prognostic indicator of adverse events in women with signs and symptoms of ischemia without obstructive CAD^12^. Whereas access to perfusion PET is limited by cost and availability, TFC is available for all patients undergoing coronary angiography. Additionally, unlike other non-invasive methods, coronary angiography has the potential to couple gold-standard epicardial CAD assessment with measures of the coronary microvasculature.

Despite the potential of TFC to be used in the diagnosis of CMVD^10, 13^, it has not been widely adopted. In this study, we establish the sex-dependent relationship between TFC and PET-derived MBF measures in patients without obstructive CAD. We show empirically that TFC correlates with MBFR, use mathematical modeling to support this surprising observation, and establish cutoff values for TFC as a diagnostic test for CMVD in women and in men.

## Methods

### Population

We identified 350 patients who underwent both perfusion PET stress testing and coronary angiography from 2012 to 2015 at the University of Pennsylvania Health System for clinical indications as part of routine medical care. We then excluded those with prior heart transplantation, obstructive CAD (defined as > 50% stenosis) on angiography, or poor image quality on coronary angiography or perfusion PET. TFC and Perfusion PET data were collected for the remaining 123 patients (age 58 +/− 12.1, 63% female). The study was approved by the University of Pennsylvania Institutional Review Board and no informed consent was required for this retrospective study using data from the Electronic Health Record.

### Rubidium-82 (82Rb) Cardiac PET Perfusion Imaging

Patients underwent a rest-dipyridamole stress 82Rb cardiac PET using a Siemens Biograph mCT PET/CT scanner. Briefly, low dose CT images were acquired for attenuation correction. Rest images were obtained with a 6-minute list-mode dynamic PET acquisition imaging while 30 mCi of 82Rb was injected intravenously as a fast bolus. Dipyridamole (0.56 mg/kg) was then administered and dynamic PET imaging was repeated with an additional 30 mCi of 82Rb three minutes after the completion of the infusion. Iterative reconstruction was performed with 2 iterations and the matrix size of 128 × 128^14, 15^. Global and regional MBF and MBFR were calculated using Syngo^®^ MBF software^16^.

### TIMI Frame Count (TFC)

Clinical coronary angiography images were used to measure TFC as outlined in Gibson et al^10^. Coronary angiograms were acquired during routine clinical care with a frame rate of 15 frames per second. Coronary angiograms were never on the same day as PET scans, but most were < 1 year apart. TFC was measured by an investigator blinded to PET parameters.

### Statistics

The relationship between TFC from different territories was analyzed using Pearson correlation coefficient (PCC). The relationship between TFC and MBF/MBFR was analyzed using linear regression analysis. Regression modeling was performed using SAS. Absolute numbers and percentages were used to describe the patient population. Continuous variables were expressed as mean +/− standard deviation. T-test was used to compare groups, and P-values<0.05 were considered significant. Adjusting MBF by heart rate-blood pressure product did not alter the results, and therefore the unadjusted MBF values are presented. Receiver operating curves (ROC) were generate and fitted using the maximum-likelihood approach^17^ implemented in the program LABROC, which is available as part of the Metz ROC Software (University of Chicago). The ROC data were fitted using a semi-parametric method, “proper” binormal model^18^, and inverse information matrix was used to estimate parameter uncertainties. Area Under the Curve (AUC) is presented and p<0.05 was considered significant.

## Results

We identified a cohort of 123 patients (age 58 +/− 12.1, 63% female) who were eligible for this study (Table 1). This population was racially diverse (54% black and 41% white) with high prevalence of cardiovascular risk factors, and similar number of patients with and without CMVD, defined as MBFR < 2.

**Table 1.**
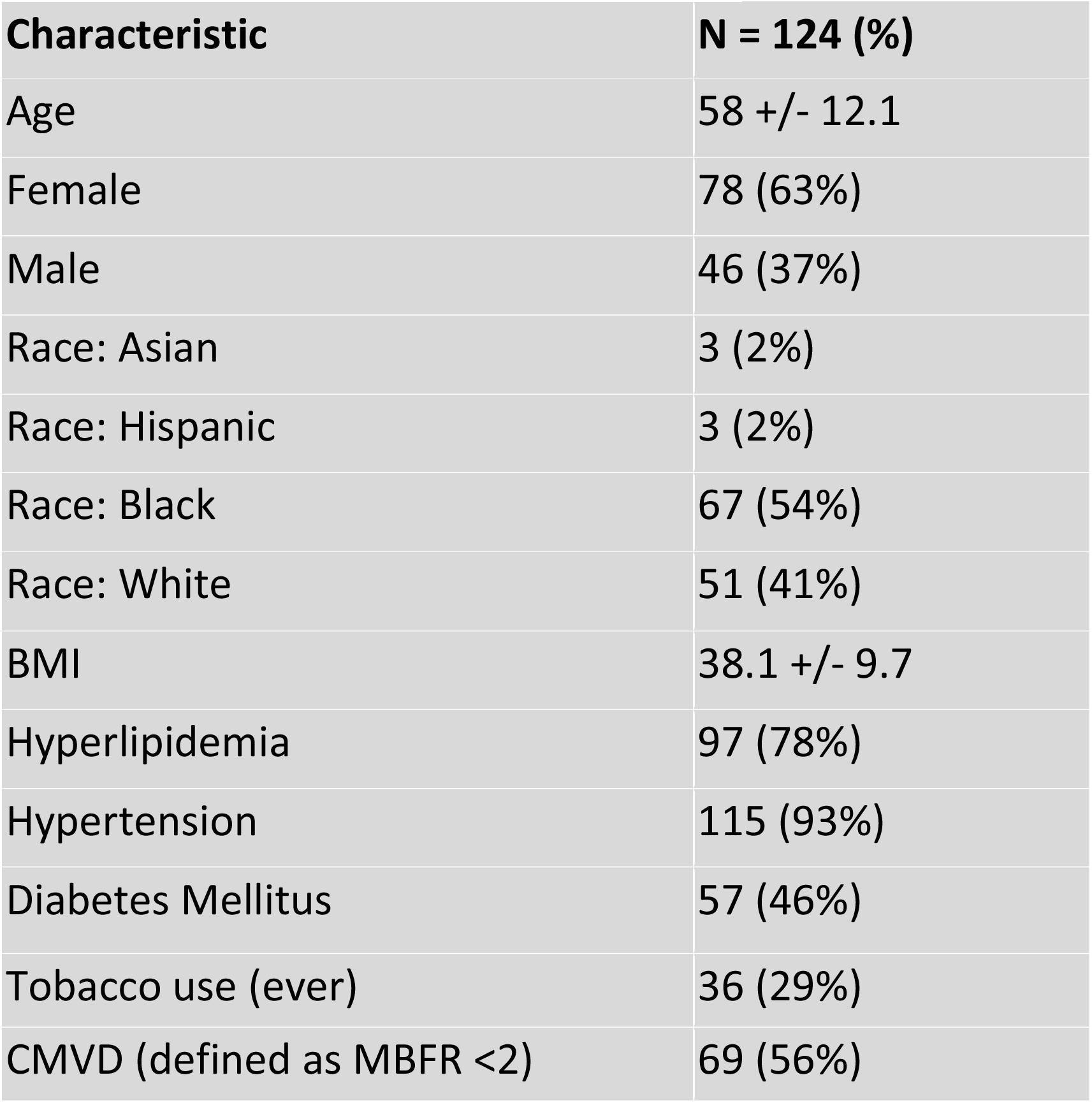
Population demographics.

| **Characteristic** | **N = 124 (%)** |
| --- | --- |
| Age | 58 +/- 12.1 |
| Female | 78 (63%) |
| Male | 46 (37%) |
| Race: Asian | 3 (2%) |
| Race: Hispanic | 3 (2%) |
| Race: Black | 67 (54%) |
| Race: White | 51 (41%) |
| BMI | 38.1 +/- 9.7 |
| Hyperlipidemia | 97 (78%) |
| Hypertension | 115 (93%) |
| Diabetes Mellitus | 57 (46%) |
| Tobacco use (ever) | 36 (29%) |
| CMVD (defined as MBFR <2) | 69 (56%) |

Because patients with CAD were excluded, there was a strong positive correlation between the global MBFR and regional MBFR for each coronary territory, with PCC 0.91, 0.96, and 0.97 for right coronary artery (RCA), left circumflex artery (LCX) and left anterior descending artery (LAD), respectively. There was a positive, although weaker, correlation in TFC between different regions (LAD vs. LCx: PCC= 0.76; LAD vs. RCA: PCC = 0.67; LCx vs. RCA: PCC = 0.65) (p<0.0001 for all territories) (Figure 1).

**Figure 1.**
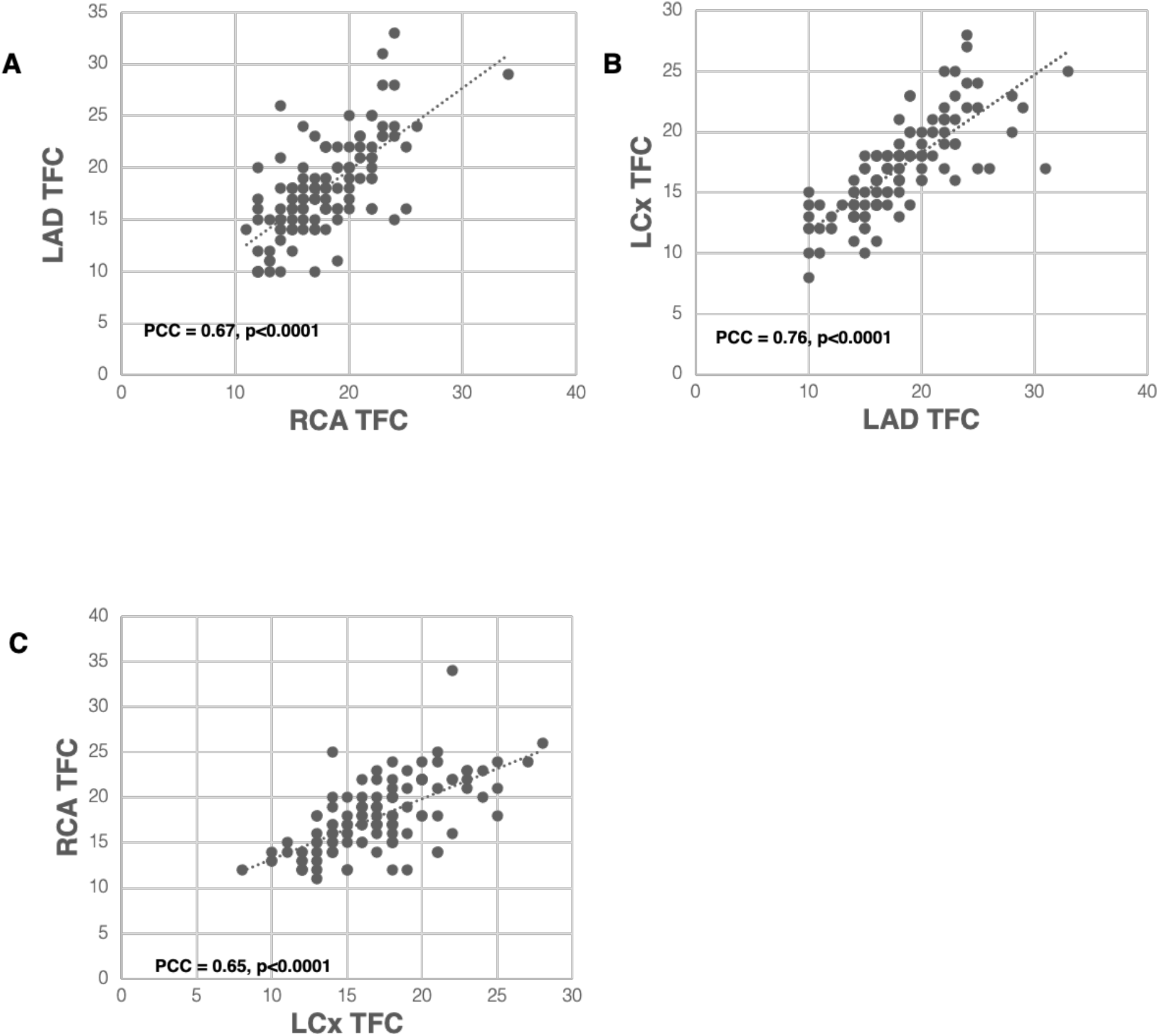
High correlation between TFC measured from different coronary territories. There is a high correlation between TFC obtained from LAD versus RCA (A, PCC=0.67), LAD versus LCX (B, PCC=0.76), and RCA versus LCX (C, PCC=0.65).

We next compared the relationship between TFC for each coronary artery and corresponding regional MBFR and found an inverse relationship between them with PCC −0.51, −0.54, and −0.51 for RCA, LCX, and LAD, respectively (Figure 2A, p < 0.0001 for all). Interestingly, there was a strong sex difference in this negative correlation between TFC and regional MBFR. In women, correlations between TFC and MBFR were −0.73, −0.74, and −0.78 for RCA, LCX, and LAD, respectively (Figure 2B, p < 0.0001 for all). The relationship between TFC and regional MBFR also existed in men but was weaker, with PCC −0.41, −0.38, and −0.35for RCA, LCX, and LAD, respectively (Figure 2C, p < 0.0001 for all).

**Figure 2.**
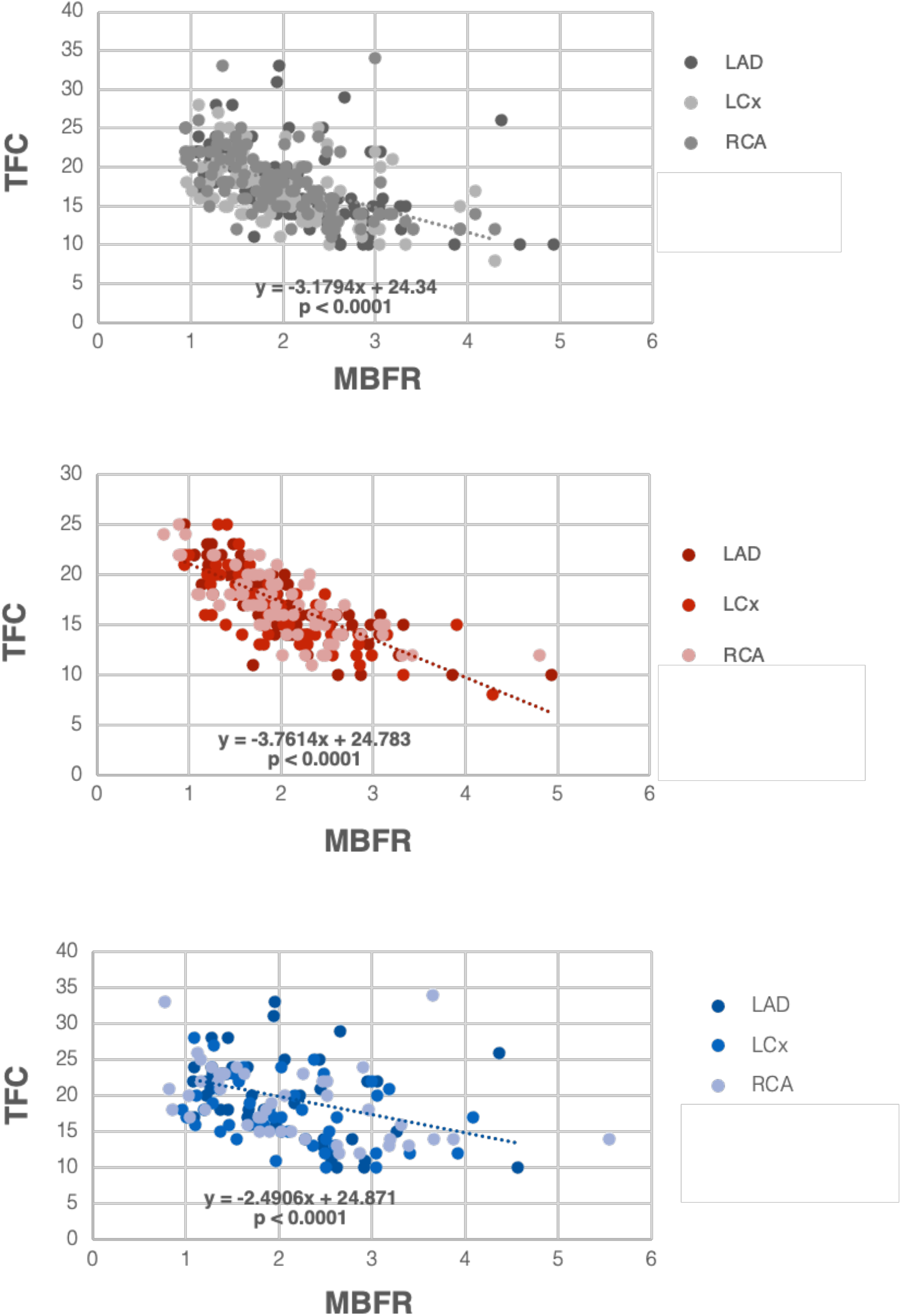
Correlation between TFC and MBFR. (A) Correlation between MBFR and TFC in all patients (p<0.0001 for all 3 coronary territories). (B) Correlation between MBFR and TFC in female patients (p<0.0001 for all 3 coronary territories). (C) Correlation between MBFR and TFC in male patients (p<0.0001 for all 3 coronary).

To gain insight into factors driving the association between TFC and MBFR, we examined the two components that comprise MBFR: rest MBF and stress MBF. There was no statistically significant correlation between TFC and rest MBF in all patients, but there was a trend towards increasing TFC with higher rest MBF in LAD and LCX. PCC between TFC and rest MBF were 0.127 (p = 0.17), −0.04 (p = 0.67), and 0.108 (p = 0.24) for LAD, RCA, and LCX, respectively (Figure 3A). There was a statistically significant, but only slight, negative relationship between TFC and stress flows. TFC versus stress MBF had correlation of −0.364, −0.512, −0.384 in LAD, RCA, and LCX, respectively (Figure 3B, p < 0.0001 for all).

**Figure 3.**
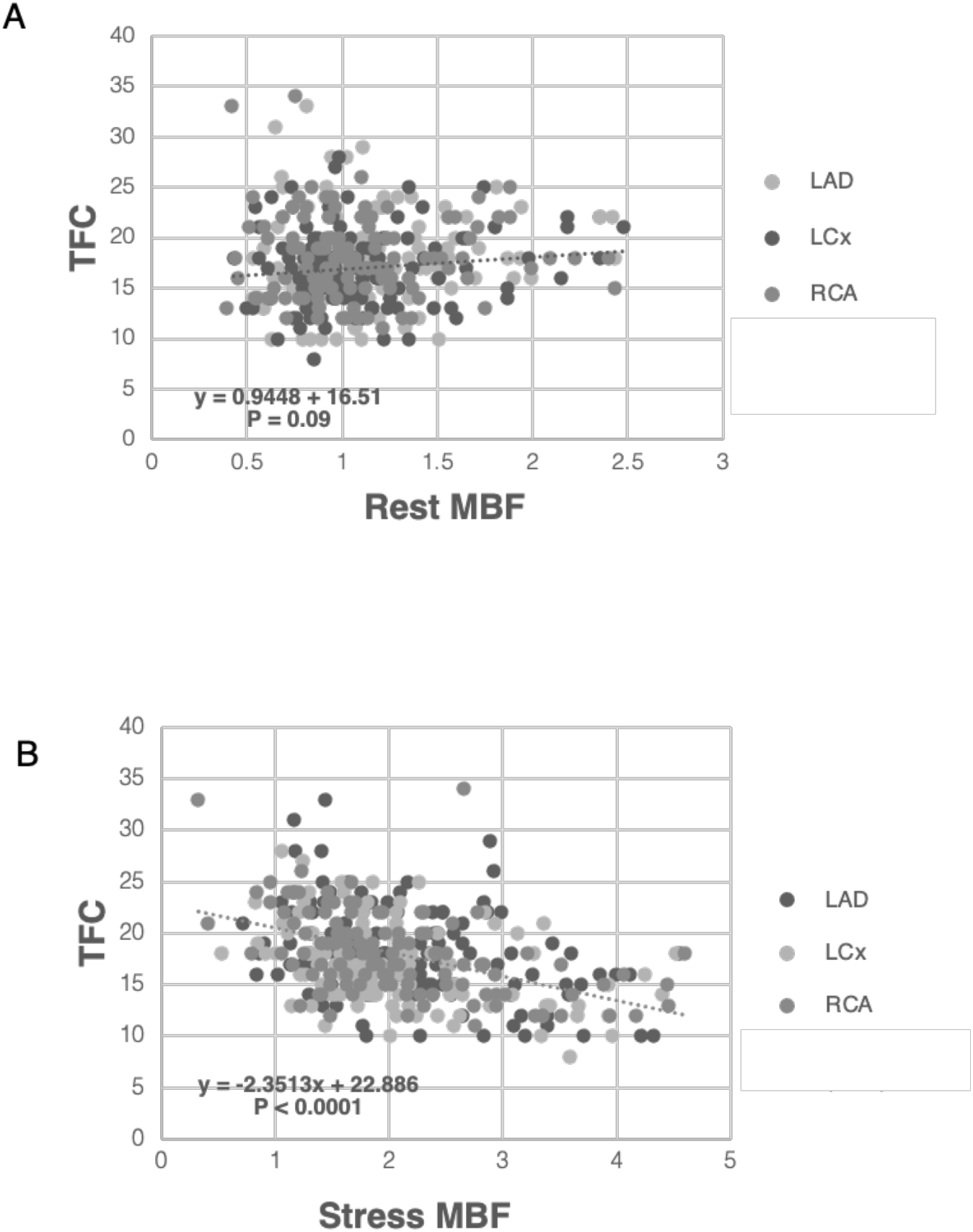
Correlation between TFC and rest and stress MBF. (A) Resting myocardial blood flow versus TFC across coronary territories in all patients lacks a significant relationship. (B) Stress myocardial blood flow versus TFC across coronary territories in all patients shows a significant but only slight inverse relationship.

TFC is a measure of how much time is required for epicardial coronary arteries to fill with contrast agent and therefore would seem to reflect coronary flow. Depending on the dose and type of sedation used during the coronary angiogram, this may represent a rest or hyperemic state. Regardless, the result that TFC correlated with MBFR, which is a unitless ratio and reflects the ability of the coronary microvasculature to vasodilate, was therefore unexpected and not intuitive. To investigate this association, we used computational modeling of the coronary arteries. First, we developed a resistance model consisting of four vascular layers: epicardial/conductive coronaries, pre-arterioles, arterioles, and capillaries (Figure 4A) and TFC was modeled as the time it took to fill the conductive vessels. The relationship between flow and diameter was preserved in capillaries, arterioles, and pre-arterioles where increased diameter led to higher flow and lower TFC (Figure 4B-4D). However, in the largest vessels, there was minimal change in flow over a large range of diameters. This is consistent with known coronary physiology where the large epicardial vessels are not the primary regulators of coronary flow. There was also a positive relationship between TFC and diameter in the largest vessels, which is in contrast to the smaller vessels.

**Figure 4.**
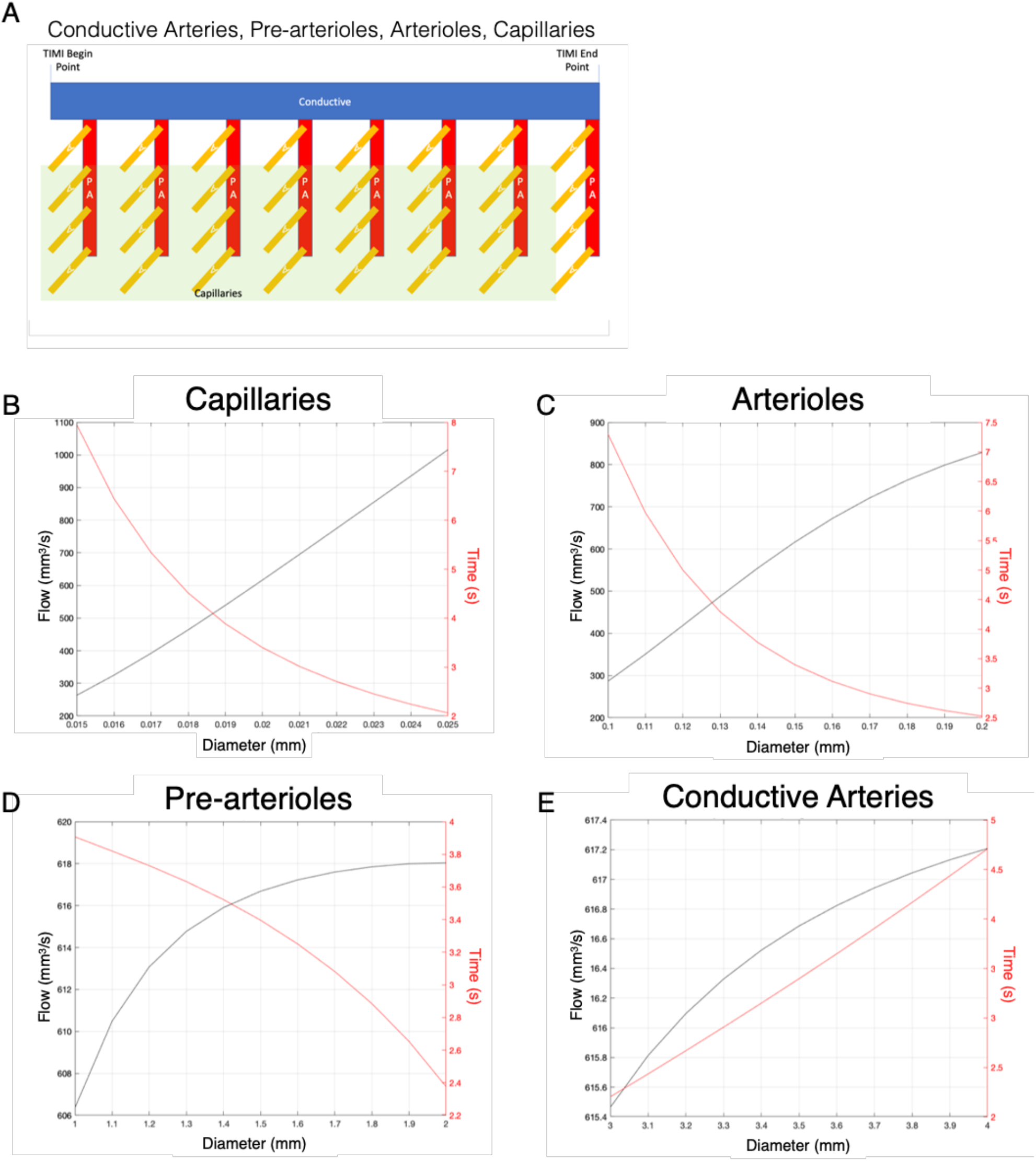
Conceptual resistance model of the coronary circulation. (A) A simplified model incorporates four vessel size compartments and defines the TFC as the time to fill the larger conductive vessel compartment (blue). (B-D) The relationship between Flow (black) and TFC measured as Time (red) with different vessel diameters highlights the expected inverse relationships between Flow and Time in capillaries (B), arterioles (C), and pre-arterioles (D). (E) Conductive arteries show minimal change in flow with large change in Time, supporting the uncoupling between resting flow and TFC seen in our cohort.

This resistance model, although conceptually useful, is limited and overlooks complex coronary branching and a wide range of vessel sizes. We therefore used a modified Zhou-Kassab-Molloi (ZKM) model, which expands the coronary tree and incorporates more accurate coronary anatomy^19^. Our implementation of this model included 24 Strahler levels (or vessel sizes), where the length and volume of each was estimated (Supplemental Figure 1A-B). As with the simplified resistance model, we found that TFC and MBFR were inversely correlated at all levels of the modeled circulation except the two largest diameter levels (Supplemental Figure 1C). Here we found that TFC and MBFR were positively associated in first order and second order vessels, which are those visualized by coronary angiography and in which TFC is measured.

To determine the potential of TFC to identify individuals with CMVD (defined as MBFR < 2), we performed ROC curve analysis for each territory separately and all together, stratified by sex (Figure 5A-D). ROC analysis showed excellent performance for TFC to identify CMVD in women with some slight variability among territories (AUC = 0.86, 0.84, and 0.89 for LAD, RCA and LCX, respectively). TFC performed less well in men in all territories (AUC = 0.75, 0.78, and 0.68 for LAD, RCA and LCX, respectively). When all territories were considered, there was a statistically significant difference in TFC performance between women and men (AUC = 0.86 vs 0.72, p = 0.003). Based on ROC curves, we established TFC thresholds that identify CMVD with sensitivity > 80% in women for all three coronary arteries (Figure 5E). The high territory-specific variability limited the sensitivity in men (Figure 5F).

**Figure 5.**
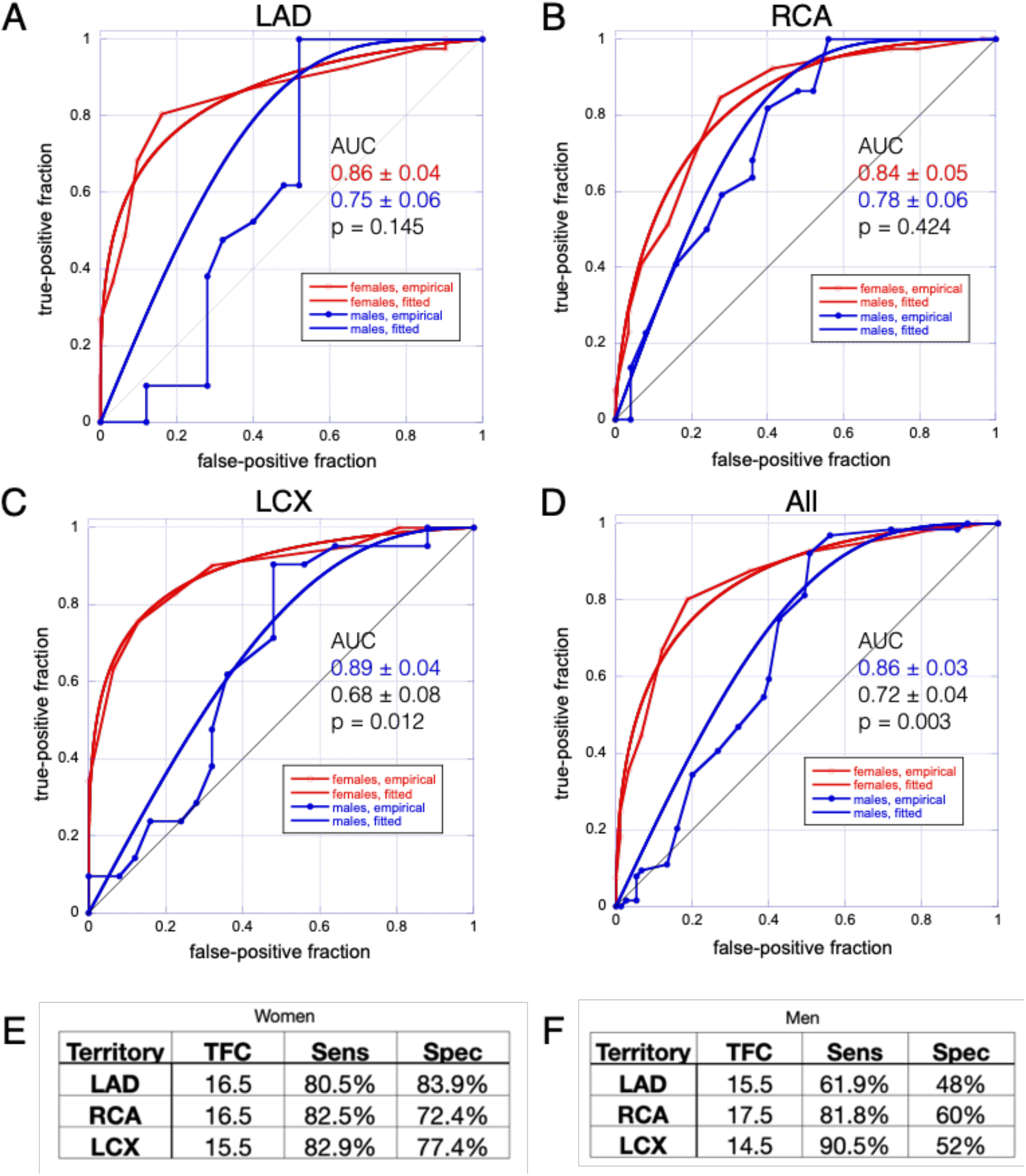
ROC analysis. (A-D) ROC analyses for TFC in LAD (A), RCA (B), LCX (C), and all territories combined (D) show that higher performances in women. (E, F), Sensitivity and specificity for TFC values chosen from ROC analyses as diagnostic thresholds for CMVD in women (E) and in men (F).

## Discussion

In a diverse cohort of patients, we establish a relationship between TFC from clinical coronary angiograms and perfusion PET MBFR and show that TFC serves as a measure of coronary microvascular *function* in women. We establish a relationship between TFC and MBFR that is stronger in women than in men. Using computational modeling, we provide a simple conceptual model then a more rigorous and anatomically realistic model to show how the relationship between vessel size, coronary flow, and TFC break down in the larger vessels that are used for TFC measurement. We test the performance of TFC in diagnosing CMVD. To our knowledge, this is the largest study to evaluate the use of TFC to diagnose CMVD in non-obstructive coronary arteries.

The strong relationship between TFC and MBFR, though counterintuitive, is supported by empiric data and computational modeling. TFC is thought to represent a measure of resting coronary flow^20^ and TFC measured before and after administration of adenosine correlate with coronary flow reserve in a cohort of 11 patients^21^. However, prior studies have shown that flow reserve is largely driven by increased resting coronary flow^22, 23^. Thus, we might expect patients with CMVD to have increased resting flows and lower TFC. In our patient cohort, there is a minimal relationship between rest MBF and TFC and a slight inverse relationship between TFC and stress MBF. Coronary modeling helps to explain these disparate data. We show that there is an uncoupling between flow and TFC in larger vessels, which allows for increased TFC in the presence of increased resting flow and decreased flow reserve. Thus, the intuitive notion that increased flow would lead to decreased time to fill is true for small vessels but not in the larger vessels where TFC is measured. That is, TFC serves as a measure of disease in these patients, rather than flow. At the patient level, this results is consistent. TFC and MBFR are both measures of CMVD, and patients with more severe CMVD have both lower MBFR and higher TFC. This is further supported by the time between PET and coronary angiography, suggesting that this reflects underlying pathology instead of a snapshot of hemodynamic parameters.

Prior work by Gibson et al. has advocated the use of TFC for assessing the coronary microvasculature in patients undergoing coronary angiography^10, 13^. Petersen et al. have shown that resting TFC independently predicts rates of hospitalization for angina in women with signs of ischemia without obstructive CAD ^24^. Our data support that these two groups of patients may have CMVD linking increased TFC with adverse outcomes. Our results further support the feasibility to quantify TFC from clinical angiograms and establish thresholds to use clinically acquired TFC as a measure of CMVD, especially in women. Given the limited availability of advanced cardiac imaging modalities and the underdiagnosis of CMVD, especially in women, widespread use of TFC during coronary angiography to inform the CMVD status may help close the gap in outcomes.

This study has a few limitations. The underlying mechanism driving the sex-difference in the relationship between TFC and MBFR is unclear. Our study cohort is enriched for women, but it is similar to prior studies that have found larger proportions of women with symptoms and non-obstructive coronaries^25^. The weaker relationship between TFC and MBFR in men therefore is likely due to greater variance rather than smaller sample size (Figure 2). Based on coronary modeling, simply having larger epicardial coronaries or more coronary vessel sizes does not account for the differences either. Sex-based differences in plaque formation and vascular remodeling have been described and may contribute to this difference ^26^. Another limitation of this work is that it is a single-center, retrospective study, with a small male population. Lastly, this study links two distinct imaging biomarkers, but further work is needed to link the heterogenous pathophysiology of CMVD with these biomarkers. MBFR and TFC both seem to reflect CMVD, however MBFR directly assesses vasodilatory capacity whereas TFC measures a single hemodynamic state.

Future studies are needed to understand sex-specific differences, to establish TFC as a diagnostic test for CMVD, and to link TFC with adverse cardiovascular outcomes in a large population without obstructive CAD. TFC has great potential as an imaging biomarker to accelerate the research into CMVD pathogenesis and may be helpful for screening patients for CMVD trials. TFC may also be a useful imaging biomarker for phenotyping CMVD in large cohorts and biobanks, such as the PennMedicine Biobank, the UK Biobank, and the Veterans Administration Million Veterans Program. However, we propose that the greatest immediate impact of this work is to further support the use of TFC as a simple, cost-effective, and widely available tool that can be applied to clinical coronary angiograms to assess for CMVD, especially in women with cardiac symptoms who may otherwise be falsely reassured by non-obstructive coronary arteries.

## Data Availability

All data produced in the present study are available upon reasonable request to the authors.

## Acknowledgments and Sources of Funding

This work was supported by NIH K08HL136890, Burroughs Wellcome Fund, and Doris Duke Charitable Foundation (MAG) and NIH R01HL149801 (MAG, SDM). This manuscript was edited at Life Science Editors.

## Figure Legends

**Supplemental Figure 1.**
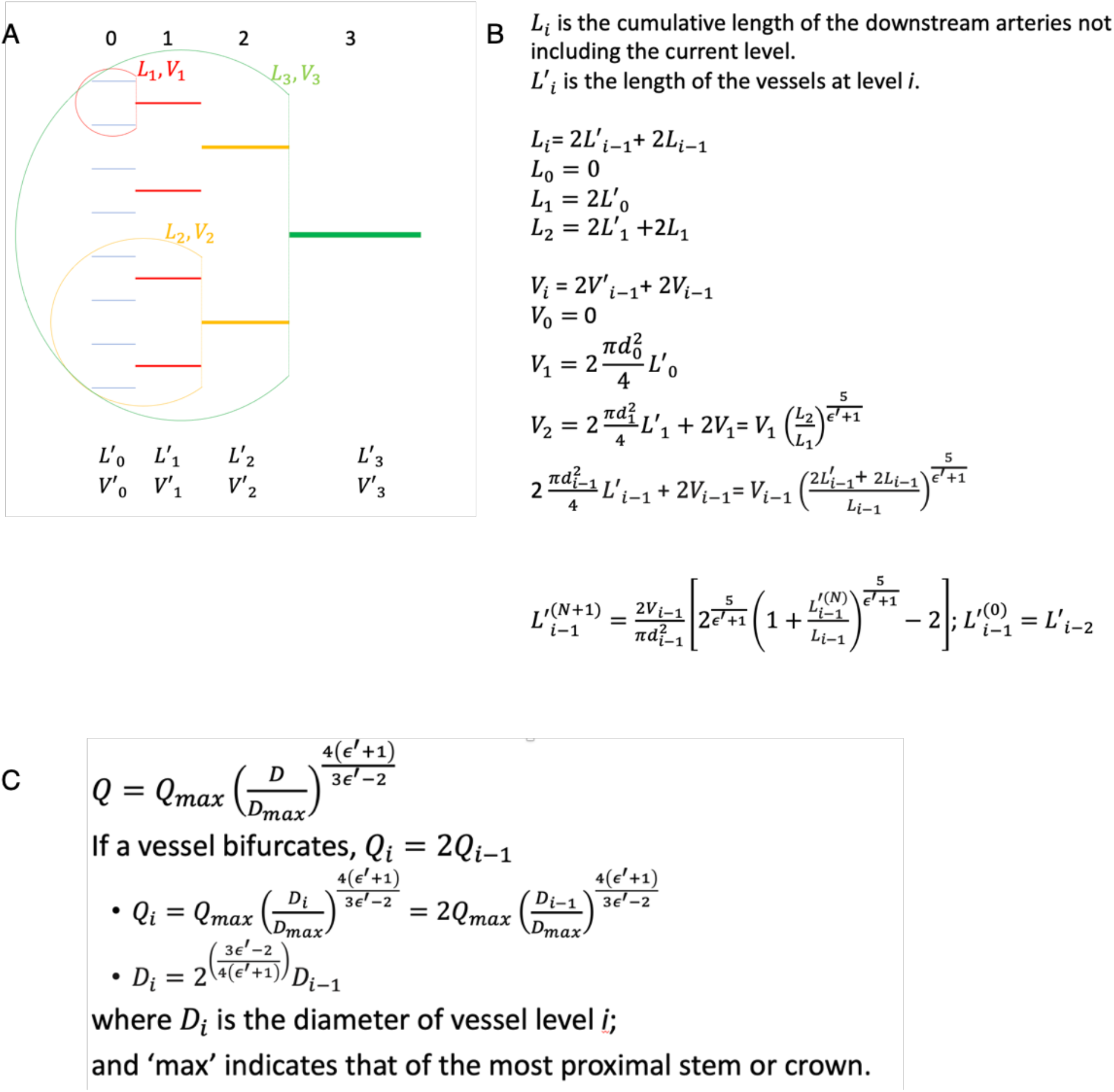
Implementation of Modified ZKM model of the coronary circulation. (A) The model is based on the stem-crown model where each vessel is a stem that branches and perfuses a territory or crown. At each level, a length (L) and Volume (V) is calculated. (B) Mathematical model outlining definitions of the L and V relative to vessel diameter (D) and with fitted fixed parameter (ε’) (C) For each level, flow (Q) is calculated as a function of D.

**Supplemental Figure 2.**
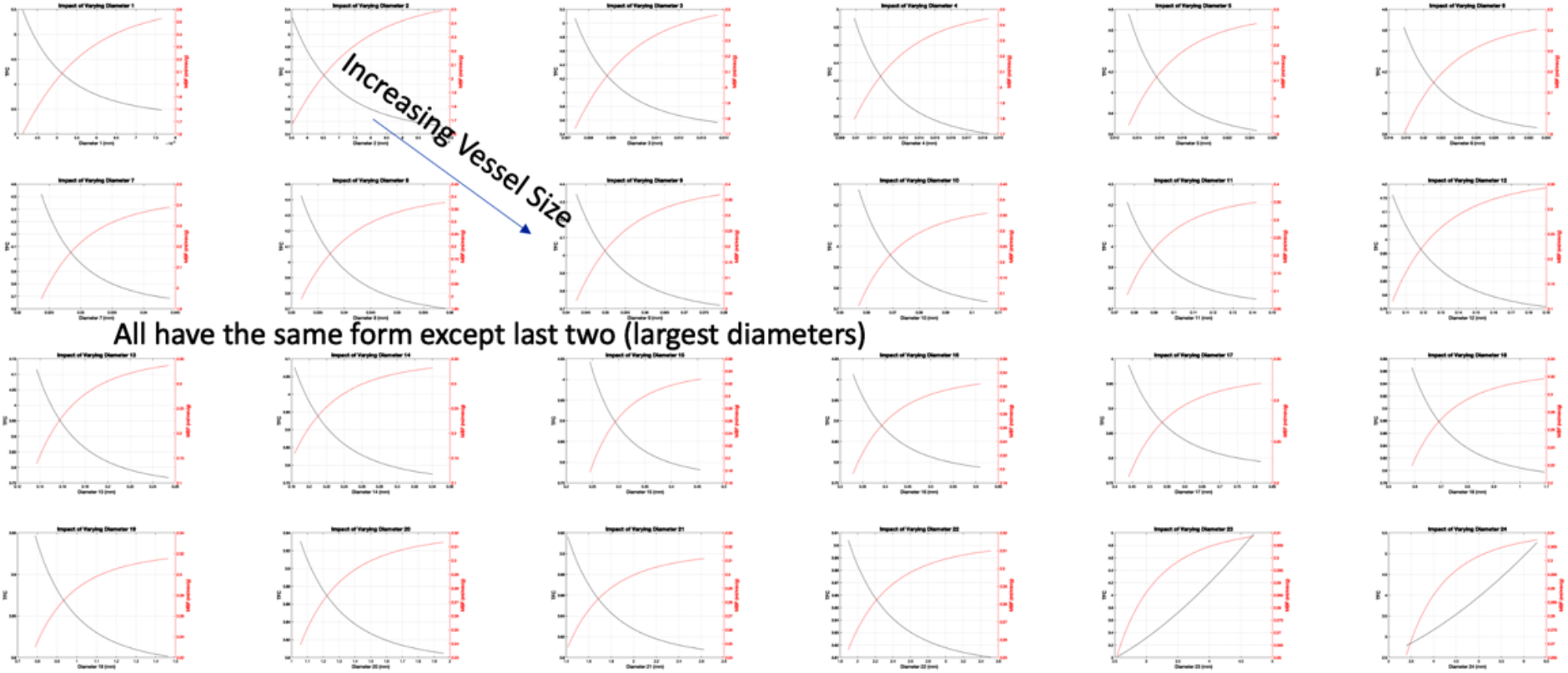
Relationship between flow and Time to fill the epicardial coronary arteries based on Modified ZKM model of the coronary circulation. There is an inverse relationship between flow (black) and time (red) in all levels of the model with the exception of the two largest levels, where there is an uncoupling of flow and time.

